# Smoking in social housing among adults in England, 2015-2020: a nationally representative survey

**DOI:** 10.1101/2022.01.11.22269062

**Authors:** Sarah E. Jackson, Hazel Cheeseman, Deborah Arnott, Robbie Titmarsh, Jamie Brown

**Affiliations:** Department of Behavioural Science and Health, University College London, London, UK; SPECTRUM Consortium, UK; Action on Smoking and Health, London, UK

**Author notes:** Corresponding author: Dr Sarah Jackson, Department of Behavioural Science and Health, University College London, 1-19 Torrington Place, London WC1E 7HB, UK.

**Keywords:** smoking, social housing, housing tenure, inequalities

## Abstract

**Objectives:** To analyse associations between living in social housing and smoking in England and evaluate progress toward reducing disparities in smoking prevalence among residents of social housing compared with other housing types.

**Design:** Nationally-representative, cross-sectional survey between January 2015 and February 2020.

**Setting:** England.

**Participants:** 105,562 adults (≥16y).

**Primary and secondary outcome measures:** Linear and logistic regression were used to analyse associations between living in social housing (vs. other housing types) and smoking status, cigarettes per day, time to first cigarette, exposure to smoking by others, motivation to stop smoking, quit attempts, and use of cessation support. Analyses adjusted for sex, age, social grade, region, and survey year.

**Results:** Adults living in social housing had twice the odds of being a smoker (OR_adj_=2.17, 95%CI 2.08- 2.27), and the decline in smoking prevalence between 2015 and 2020 was less pronounced in this high-risk group (−7%; OR_adj_=0.98, 95%CI 0.96-1.01) than among adults living in other housing types (− 24%; OR_adj_=0.95, 95%CI 0.94-0.96; housing tenure*survey year interaction *p*=0.020). Smokers living in social housing were more addicted than those in other housing (smoking within 30 minutes of waking: OR_adj_=1.50, 95%CI 1.39-1.61), but were no less motivated to stop smoking (OR_adj_=1.06, 95%CI 0.96-1.17) and had higher odds of having made a serious attempt to quit in the past year (OR_adj_=1.16, 95%CI 1.07-1.25). Among smokers who had tried to quit, those living in social housing had higher odds of using evidence-based cessation support (OR_adj_=1.22, 95%CI 1.07-1.39) but lower odds of remaining abstinent (OR_adj_=0.63, 95%CI 0.52-0.76).

**Conclusions:** There remain stark inequalities in smoking and quitting behaviour by housing tenure in England, with declines in prevalence stalling between 2015 and 2020 despite progress in the rest of the population. In the absence of targeted interventions to boost quitting among social housing residents, inequalities in health are likely to worsen.

**Strengths and limitations of this study:** A major strength of this study was the large sample, which was representative of adults living in England.

Another strength was the broad range of smoking outcomes assessed, offering a detailed view of smoking behaviour among people living in social housing compared with those living in other housing types.

The main limitation was that all outcomes were self-reported, introducing scope for bias.

## Introduction

Tobacco smoking is one of the leading drivers of health inequalities in England (1). Higher smoking prevalence is associated with almost every indicator of socioeconomic disadvantage (2) and progress to reduce smoking prevalence has historically been slower among disadvantaged groups (3,4). Understanding and alleviating this inequality is a priority for public health research and policy.

Housing tenure is an indicator of socioeconomic position that is particularly strongly linked with smoking (5). A large survey in England in 2015-17 revealed 34% of adults living in social housing were smokers, compared with 15% of people living in other housing types (e.g. home owners or private renters) (6). Strikingly, smokers living in social housing were no less motivated to quit, but were only around half as likely to be successful when they tried (6). This report prompted calls for targeted action to address this disparity (7). The UK Government’s 2017 tobacco control plan for England committed to eliminating inequalities and reducing smoking prevalence in groups with the highest rates (8). More recently, the Government committed to ‘levelling up’ disparities in health outcomes, incomes, and educational opportunities (9). What, if any, subsequent progress has been made in tackling smoking in social housing is unclear.

Using data from a nationally-representative survey of more than 100,000 adults in England between 2015 and 2020, this study aimed to provide an update on smoking in social housing in England and evaluate progress toward reducing disparities in smoking prevalence among residents of social housing compared with other housing types.

## Method

### Design and population

This was a cross-sectional national survey of a representative sample of adults in England. Data on housing tenure, smoking, and smoking cessation were collected in the Smoking Toolkit Study between January 2015 and February 2020 [23]. Data on housing tenure have not been collected since the Covid-19 pandemic required data collection to move from face-to-face to telephone interviews in March 2020, so these are the most up-to-date data available.

The Smoking Toolkit Study uses a hybrid of random probability and simple quota sampling to select a new sample of approximately 1,700 adults aged ≥16 years each month. Full details of the study’s methods are available elsewhere, and comparisons with national data and cigarette sales indicate that key variables such as sociodemographic characteristics and smoking prevalence are nationally representative (10,11).

### Patient and public involvement

The wider toolkit study has been discussed with a diverse patient and public involvement (PPI) group, and the authors regularly attend and present at meetings at which patients and public are included. Interaction and discussion at these events help to shape the broad research priorities and questions. There is also a mechanism for generalised input from the wider public: each month interviewers seek feedback on the questions from all 1,700 respondents, who are representative of the English population. This feedback is limited, and usually simply relates to understanding of questions and item options. No patients or members of the public were involved in setting the research questions or the outcome measures, nor were they involved in the design and implementation of this specific study. There are no plans to involve patients in dissemination.

### Measures

Housing tenure was categorised as ‘social housing’ (homes belonging to a housing association or rented from local authority; coded 1) vs. ‘other housing’ (homes bought on a mortgage, owned outright, rented from private landlord, or other; coded 0).

The smoking outcomes examined were:

i. *among all adults*: cigarette smoking prevalence;
ii. *among current smokers*: mean cigarettes per day (CPD) and percentage who smoke within 30 minutes of waking (as markers of cigarette dependence), high motivation to stop (‘really want and plan to stop within 3 months’ (12)), and regular exposure to smoking by others;
iii. *among past-year smokers*: percentage with a past-year quit attempt; and
iv. *among smokers with quit attempts in the past year*: percentage not currently smoking, and who used cessation support (behavioural, nicotine replacement therapy (NRT) over the counter (OTC), electronic cigarettes (e-cigarettes), or prescription medication).

Covariates were sex, age, occupational social grade (assessed using the National Readership Survey classification (13)), government office region, and survey year.

### Statistical analysis

Data were analysed using SPSS V.27. Variables were weighted using rim (marginal) weighting to match an English population profile relevant to the time each monthly survey was conducted on the dimensions of age, social grade, region, housing tenure, ethnicity and working status within sex derived from English census data, ONS mid-year estimates and other random probability surveys (10). Missing data were removed on a per-analysis basis for each outcome.

We used linear regression (continuous outcomes) and logistic regression (binary outcomes) models to analyse associations between housing tenure (social housing vs. other housing) and smoking outcomes, with and without adjustment for covariates. To test whether the effectiveness of use of evidence-based support for cessation differed by housing tenure, accounting for differences in dependence, we used logistic regression to test the interaction between housing tenure and use of evidence-based support, adjusting for covariates and measures of dependence (cigarettes per day and smoking within 30 minutes of waking).

To examine differences in smoking prevalence trends by housing tenure over the study period, we graphically displayed annual data and reran the adjusted logistic regression model for smoking prevalence adding the interaction term between housing tenure and survey year (modelled as a continuous variable). We then ran stratified analyses in which the association between smoking prevalence and survey year was tested separately for each housing type (social vs. other) to provide more information as to the nature of the difference between groups.

## Results

A total of 105,562 adults aged ≥16 years responded to the Smoking Toolkit Study survey between January 2015 and February 2020. Sample characteristics are shown in Table 1.

**Table 1.**
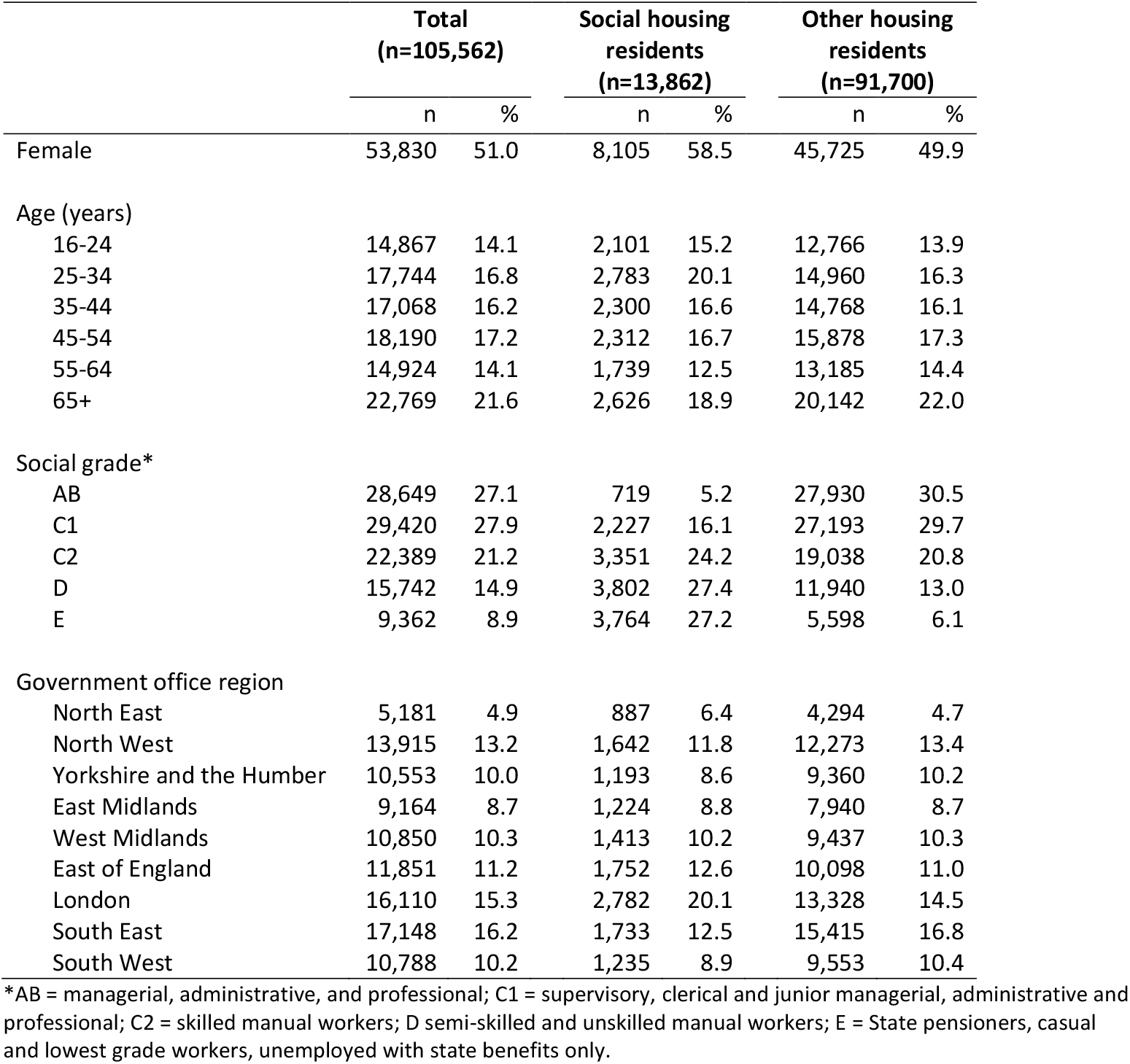
Sample characteristics

|  | **Total**<br>**(n=105,562)** |  | **Social housing**<br>**residents**<br>**(n=13,862)** |  | **Other housing**<br>**residents**<br>**(n=91,700)** |  |
| --- | --- | --- | --- | --- | --- | --- |
|  | n | % | n | % | n | % |
| Female | 53,830 | 51.0 | 8,105 | 58.5 | 45,725 | 49.9 |
| Age (years) |  |  |  |  |  |  |
| 16-24 | 14,867 | 14.1 | 2,101 | 15.2 | 12,766 | 13.9 |
| 25-34 | 17,744 | 16.8 | 2,783 | 20.1 | 14,960 | 16.3 |
| 35-44 | 17,068 | 16.2 | 2,300 | 16.6 | 14,768 | 16.1 |
| 45-54 | 18,190 | 17.2 | 2,312 | 16.7 | 15,878 | 17.3 |
| 55-64 | 14,924 | 14.1 | 1,739 | 12.5 | 13,185 | 14.4 |
| 65+ | 22,769 | 21.6 | 2,626 | 18.9 | 20,142 | 22.0 |
| Social grade* |  |  |  |  |  |  |
| AB | 28,649 | 27.1 | 719 | 5.2 | 27,930 | 30.5 |
| C1 | 29,420 | 27.9 | 2,227 | 16.1 | 27,193 | 29.7 |
| C2 | 22,389 | 21.2 | 3,351 | 24.2 | 19,038 | 20.8 |
| D | 15,742 | 14.9 | 3,802 | 27.4 | 11,940 | 13.0 |
| E | 9,362 | 8.9 | 3,764 | 27.2 | 5,598 | 6.1 |
| Government office region |  |  |  |  |  |  |
| North East | 5,181 | 4.9 | 887 | 6.4 | 4,294 | 4.7 |
| North West | 13,915 | 13.2 | 1,642 | 11.8 | 12,273 | 13.4 |
| Yorkshire and the Humber | 10,553 | 10.0 | 1,193 | 8.6 | 9,360 | 10.2 |
| East Midlands | 9,164 | 8.7 | 1,224 | 8.8 | 7,940 | 8.7 |
| West Midlands | 10,850 | 10.3 | 1,413 | 10.2 | 9,437 | 10.3 |
| East of England | 11,851 | 11.2 | 1,752 | 12.6 | 10,098 | 11.0 |
| London | 16,110 | 15.3 | 2,782 | 20.1 | 13,328 | 14.5 |
| South East | 17,148 | 16.2 | 1,733 | 12.5 | 15,415 | 16.8 |
| South West | 10,788 | 10.2 | 1,235 | 8.9 | 9,553 | 10.4 |

\*AB = managerial, administrative, and professional; C1 = supervisory, clerical and junior managerial, administrative and professional; C2 = skilled manual workers; D semi-skilled and unskilled manual workers; E = State pensioners, casual and lowest grade workers, unemployed with state benefits only.

Associations between housing tenure and smoking outcomes are shown in Table 2. After adjustment for sex, age, social grade, region, and survey year, adults living in social housing had more than double the odds of being a smoker compared with those living in other housing types. Current smokers living in social housing smoked on average one more cigarette per day and had 50% higher odds of smoking their first cigarette of the day within 30 minutes of waking, indicating significantly higher levels of addiction. Their level of motivation to stop smoking did not differ significantly from those living in other housing types, nor did the odds of reporting regular exposure to smoking by others. Smokers living in social housing had 16% higher odds of having made a serious attempt to quit in the past year than those living in other housing types. Among smokers who had tried to quit in the past year, those living in social housing had 22% higher odds of using evidence-based cessation support (specifically, e-cigarettes or prescription medication) but 37% lower odds of remaining abstinent. This does not mean evidence-based cessation support was less effective for smokers living in social housing: after adjustment for level of dependence, the association between use of evidence-based support and cessation did not differ significantly by housing tenure (interaction OR_adj_ 0.93, 95% CI 0.64-1.34, *p*=0.684).

**Table 2.** Smoking and cessation behaviour in social housing compared to other housing, January 2015 to February 2020 (n=105,616)

|  | Social housing | Other housing | Unadjusted |  |  | Adjusted** |  |  |
| --- | --- | --- | --- | --- | --- | --- | --- | --- |
|  |  |  | OR/B* | 95% CI | p | OR/B | 95% CI | p |
| *All adults* | n=13,862 | n=91,700 |  |  |  |  |  |  |
| % Cigarette smokers | 33.5 | 14.8 | 2.91 | 2.80-3.03 | <0.001 | 2.17 | 2.08-2.27 | <0.001 |
| *Current cigarette smokers* | n=4,637 | n=13,525 |  |  |  |  |  |  |
| Mean cigarettes per day | 12.2 | 10.5 | 1.72 | 1.45-1.99 | <0.001 | 0.99 | 0.71-1.27 | <0.001 |
| % First smoke within 30 min of waking | 57.4 | 42.6 | 1.82 | 1.70-1.94 | <0.001 | 1.50 | 1.39-1.61 | <0.001 |
| % High motivation to stop | 14.7 | 15.0 | 0.97 | 0.89-1.07 | 0.575 | 1.06 | 0.96-1.17 | 0.284 |
| % Regular exposure to smoking by others | 68.4 | 68.6 | 0.99 | 0.92-1.06 | 0.778 | 1.01 | 0.94-1.10 | 0.749 |
| *Past-year smokers* | n=4,923 | n=15,054 |  |  |  |  |  |  |
| % Past year quit attempt | 32.4 | 30.9 | 1.07 | 1.00-1.15 | 0.054 | 1.16 | 1.07-1.25 | <0.001 |
| *Past year quit attempt* | n=1,551 | n=4,530 |  |  |  |  |  |  |
| % Not currently smoking | 11.6 | 18.9 | 0.56 | 0.47-0.67 | <0.001 | 0.63 | 0.52-0.76 | <0.001 |
| % Used any cessation support*** | 59.0 | 54.4 | 1.20 | 1.07-1.35 | 0.002 | 1.22 | 1.07-1.39 | 0.003 |
| % Used behavioural support | 2.8 | 2.2 | 1.25 | 0.87-1.80 | 0.229 | 1.20 | 0.80-1.80 | 0.377 |
| % Used NRT OTC | 13.4 | 13.0 | 1.04 | 0.88-1.23 | 0.671 | 0.88 | 0.73-1.07 | 0.189 |
| % Used e-cigarettes | 33.9 | 32.1 | 1.08 | 0.96-1.23 | 0.196 | 1.19 | 1.04-1.36 | 0.012 |
| % Used prescription medication | 9.0 | 7.1 | 1.28 | 1.04-1.58 | 0.020 | 1.33 | 1.05-1.68 | 0.017 |

\*B can be interpreted as the mean (unadjusted/adjusted, as relevant) difference between the social housing and other housing groups. \*\*OR/B adjusted for sex, age, social grade, government office region, and survey year. \*\*\*Any cessation support includes behavioural support, nicotine replacement therapy (NRT) bought over-the-counter (OTC), e-cigarettes, and prescription medication.
Number of missing cases per variable: % cigarette smokers n=51 (0.0%); mean cigarettes per day n=325 (1.8%); % first smoke within 30 min of waking n=81 (0.4%); % high motivation to stop n=33 (0.2%); % regular exposure to smoking by others n=0 (0.0%); % past year quit attempt n=556 (2.8%); % not currently smoking n=0 (0.0%); % used cessation support n=0 (0.0%).

Figure 1 shows annual smoking prevalence estimates over the study period. There was a significant interaction between housing tenure and survey year on smoking prevalence (OR_adj_ 1.03, 95% CI 1.01-1.06, *p*=0.020). Stratified analyses showed that there was a significant linear decline in smoking prevalence between 2015 and 2020 among adults living in other housing types (OR_adj_ 0.95, 95% CI 0.94-0.96, *p*<0.001), with prevalence falling by 24% (from 16.0% in 2015 to 12.1% in 2020). However, the decline among adults living in social housing over the same period was not statistically significant (OR_adj_ 0.98, 95% CI 0.96-1.01, *p*=0.120), falling by just 7% (from 35.3% in 2015 to 32.7% in 2020).

**Figure 1.**
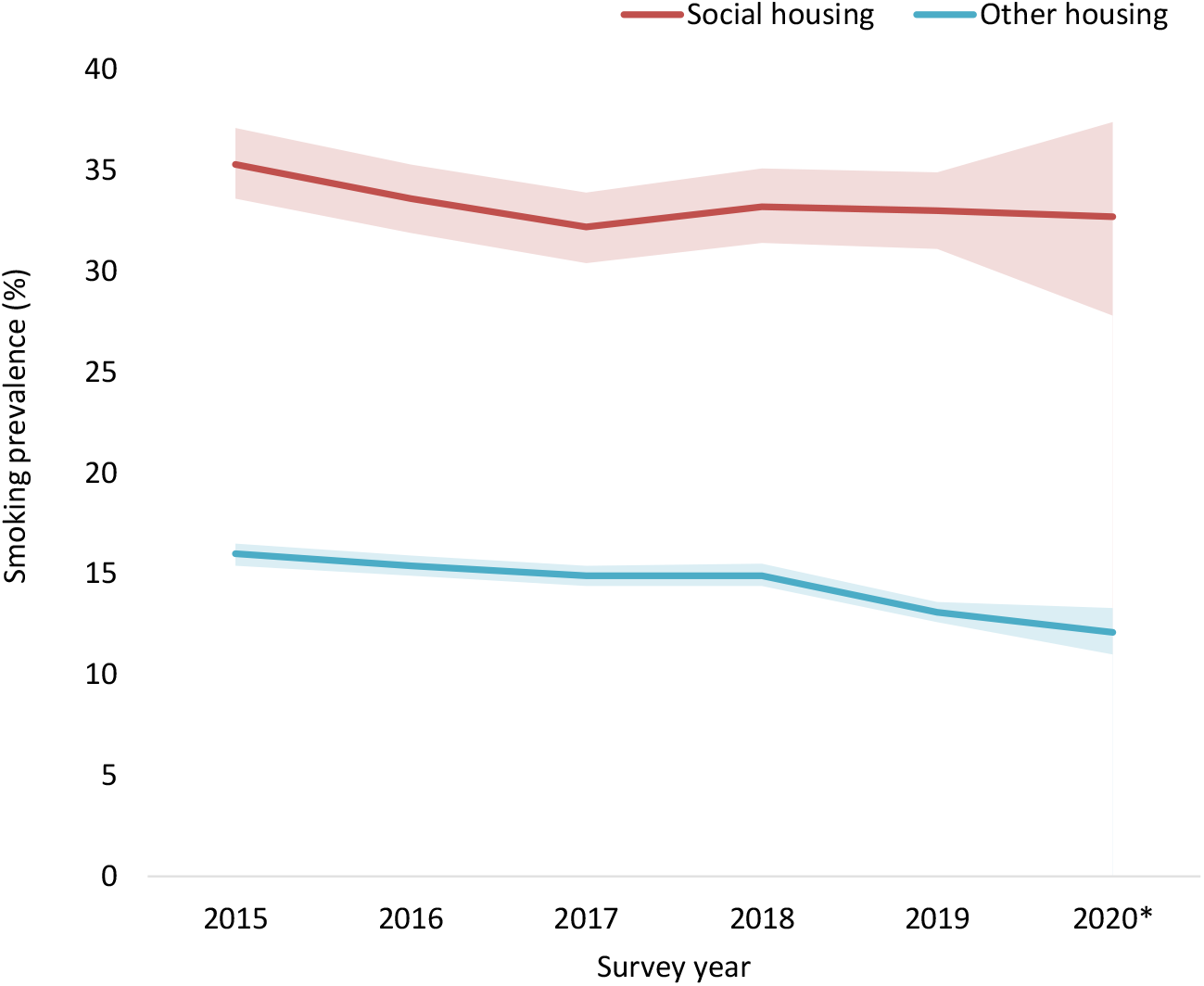
Annual smoking prevalence among adults in England living in social housing compared with other housing tenures, January 2015 through February 2020. Shaded bands indicate 95% confidence intervals. Bases (weighted n): social housing 2015 n=2849, 2016 n=2910, 2017 n=2717, 2018 n=2579, 2019 n=2420, 2020 n=373; other housing 2015 n=17132, 2016 n=17520, 2017 n=17662, 2018 n=18106, 2019 n=18215, 2020 n=3029. *Note: Data for 2020 are from January and February only.

## Discussion

This study extends the existing evidence base on smoking in social housing in England. Results showed adults who live in social housing remain more likely to smoke, and the general decline in smoking prevalence over recent years has stalled in this high-risk group compared with adults living in other housing types, indicating worsening inequalities in smoking on this measure. While smokers living in social housing are more addicted than those living in other housing, they are equally motivated to quit, more likely to make a quit attempt, and more likely to use support. Yet they are less likely to be successful in stopping.

The results are consistent with those of a previous analysis that included data from 2015-17 (6), suggesting there has been little change in smoking inequalities between adults who live in social versus other types of housing over recent years. The only notable difference was that in this analysis, use of prescription medication as a cessation aid was significantly higher among smokers living in social housing than other housing types when it had not been previously. This could be explained by a smaller reduction in use of prescription medication from the original to current analysis among smokers living in social housing (from 9.3% to 9.0%) than those living in other housing types (from 8.2% to 7.1%). It is encouraging that smokers in social housing were more likely to access evidence-based support, which can substantially increase their chances of quitting successfully, because their higher levels of dependence and various social and environmental barriers make it more difficult for them to successfully stop smoking. However, with four in ten quitters not using any form of evidence-based support, there remains room for improvement in helping smokers in social housing (and other housing tenures) to access effective support and translate more quit attempts into long-term cessation.

Without targeted action, smoking-related disparities are likely to have significant implications for the health of people and their families living in social housing. The adverse effects of smoking on health and life expectancy are well established, and the transmission to the next generation (14), but much of the harm caused by smoking can be reversed by quitting (15,16). This offers huge policy potential to ‘level up’ and reduce the damage smoking causes. Various approaches have been suggested to better support smokers in social housing, including ways in which social landlords can maximise their opportunity to improve tenants’ wellbeing (7). Most recently, the All Party Parliamentary Group on Smoking and Health recommended an at-scale intervention to provide free e-cigarettes and behavioural support to smokers in social housing (17) based on a successful pilot in Salford in the North of England (18).

A major strength of this study was the large, representative sample. The main limitation was that all outcomes were self-reported, introducing scope for bias. Measurement of quit attempts and use of support relied on recall of the past year and quit success was not biochemically verified. While the latter would be a significant limitation in randomised trials (because smokers who receive active treatment may feel social pressure to claim abstinence) social pressure and the associated rate of misreporting is low in population surveys (19). Moreover, we would not expect the extent of misreporting to differ by housing tenure meaning our results are unlikely to materially be affected.

In conclusion, there remain stark inequalities in smoking and quitting behaviour by housing tenure in England, with declines in prevalence stalling between 2015 and 2020 despite progress in the rest of the population. In the absence of targeted interventions to boost quitting among social housing residents, inequalities in health are likely to worsen. In the context of the UK Government’s commitment to levelling up, tackling smoking in social housing should be an urgent priority.

## Data Availability

Data are available on request from the corresponding author.

## Declarations

## Competing interests

JB has received unrestricted research funding from Pfizer, who manufacture smoking cessation medications. All authors declare no financial links with tobacco companies or e-cigarette manufacturers or their representatives.

## Funding

This work was supported by Cancer Research UK (C1417/A22962).

## Author contributions

All authors conceived and designed the study. SJ analysed the data and wrote the first draft. All authors provided critical revisions.

## Ethical approval

Ethical approval for the STS was granted by the UCL Ethics Committee (ID 0498/001). The data are not collected by UCL and are anonymised when received by UCL.

## Data sharing

Data are available on request from the corresponding author.

